# The Clinical Characteristics and Prognostic Factors of Acute Graft-Versus-Host Disease in Stem Cell Transplant Patients Treated with Cyclosporine

**DOI:** 10.1101/2025.11.03.25339437

**Authors:** Xi-Bin Wang, Zi-Zhong Lin, Jie-Mei Chen, Dao-Hai Cheng, Fu-Li Qin, Tao-Tao Liu

## Abstract

**Background:** Calcium-modulated inhibitors are commonly used after allogeneic haematopoietic stem cell transplantation (allo-HSCT), and the risk factors for the development of acute graft-versus-host disease (aGVHD) may differ among immunosuppressive regimens, presenting a challenge for clinical prediction. Cyclosporine (CsA) is a frequently used immunosuppressant in the postoperative period. By investigating the clinical characteristics and risk factors of aGVHD in allo-HSCT treated with the CsA regimen, we aspire to furnish evidence-based proof for patients applying CsA after allo-HSCT.

**Methods:** This retrospective study based on the clinical data of patients who received allo-HSCT and used CsA in the First Affiliated Hospital of Guangxi Medical University from June 2020 to July 2023, calculated the incidence rate of aGVHD, and analyzed the risk factors of poor prognosis through logical regression. Simultaneously perform ROC curve analysis.

**Results:** A total of 129 subjects were enrolled in this study, among whom 19 patients developed aGVHD. The sites of aGVHD involvement were skin in 11 cases (57.9%) and intestine in 8 cases (42.1%). Univariate analysis results showed that CsA concentration on days 15 ~ 21 after transplantation, bacterial infection before transplantation, fungal infection before transplantation, and pretreatment programme were the associated factors with aGVHD. Multifactorial analysis revealed that CsA concentration on days 15~21 after transplantation (OR=0.987, 95% CI=0.978~0.997, *P*=0.011), CsA dose-normalized concentration on days 15 ~ 21 after transplantation (OR=1.015, 95% CI=1.001 ~ 1.029, *P*=0.035), bacterial infection before transplantation (OR=0.255, 95%CI=0.079 ~ 0.823, *P*=0.022), and pretreatment programme (OR=0.488, 95% CI=0.252 ~ 0.944, *P*=0.033) were independent risk factors for aGVHD. After transplantation, the ideal threshold of CsA concentration is 136.45 ng/ml.

**Conclusion:** This study highlights the necessity for patients undergoing allo-HSCT and receiving CsA regimens to be particularly vigilant about the pretreatment programme, the monitoring of CsA or CsA dose-normalized concentration on days 15 ~ 21 after transplantation, and to avoid bacterial infections before surgery.

## Introduction

In the treatment of malignant and non-malignant haematological disorders, allogeneic haematopoietic stem cell transplantation (allo-HSCT) is often used ^[1]^. However, acute graft-versus-host disease (aGVHD) poses a major complication after allo-HSCT. It is said that around 30~50% of transplanted patients will experience aGVHD, and 14% of them will have severe (grade III~IV) aGVHD, which will affect patient survival and prognosis^[2]^. It is the main reason for post-operative mortality in allo-HSCT^[3]^. aGVHD commonly emerges within the initial 100 days subsequent to allo-HSCT, during this time, the immunoreactive cells present in the transplanted tissue activate T cells, resulting in an augmentation of their numbers and facilitating their differentiation, this activation triggers a sequence of immune reactions that mainly focus on the skin, gastrointestinal tract, and liver, leading to the typical symptoms related to aGVHD. It is demonstrated that the crucial elements in the progression of aGVHD after allo-HSCT are associated with imbalance of the intestinal microflora^[3]^, the donor and recipient’s ages, the types of hematological diseases, and the intensity of the conditioning regimen ^[4]^. One study employed a machine learning method to predict aGVHD, aiming to predict the occurrence of aGVHD based on risk assessment^[5]^. However, many biomarkers in the model are controversial, primarily because of the complex factors affecting laboratory physicochemical properties. Moreover, the model’s computational complexity limits its dissemination and application.

After allo-HSCT, the calcineurin inhibitor CsA is typically used as the primary regimen for the prevention of aGVHD. Due to significant pharmacokinetic variability among individuals, therapeutic drug monitoring of CsA is usually performed. However, whether CsA concentration should be considered a risk factor for aGVHD has not yet been reported, which warrants further investigation. In this study, we explored the risk factors for the occurrence of aGVHD by conducting a multifactorial analysis of clinical indicators in allo-HSCT patients in conjunction with CsA drug concentrations. Our goal is to enhance the efficacy of allo-HSCT transplantation.

## Patients and methods

### Research Object

We enrolled 129 patients with hematological diseases who received Allo-HSCT at the the First Affiliated Hospital of Guangxi Medical University’s Haematopoietic Stem Cell Transplantation Centre from June 2020 to July 2023. CsA, at a certain dose or frequency, had been used before and after the Allo-HSCT. The inclusion criteria are: meeting the diagnostic criteria of each primary species and undergoing Allo-HSCT for the first time, studies are also being conducted on the use of CsA regimens for therapy in cases involving mismatched related donors (MMRD) and unrelated donors (UD). This study was approved by the Ethics Committee of the First Affiliated Hospital of GuangXi Medical University(NO. 2024-E748-01), and conducted in accordance with the principles outlined in the tenets of the Declaration of Helsinki. Informed consent was waived due to the retrospective nature of this study. The basic information of the patients was presented in Table 1.

**Table 1.**
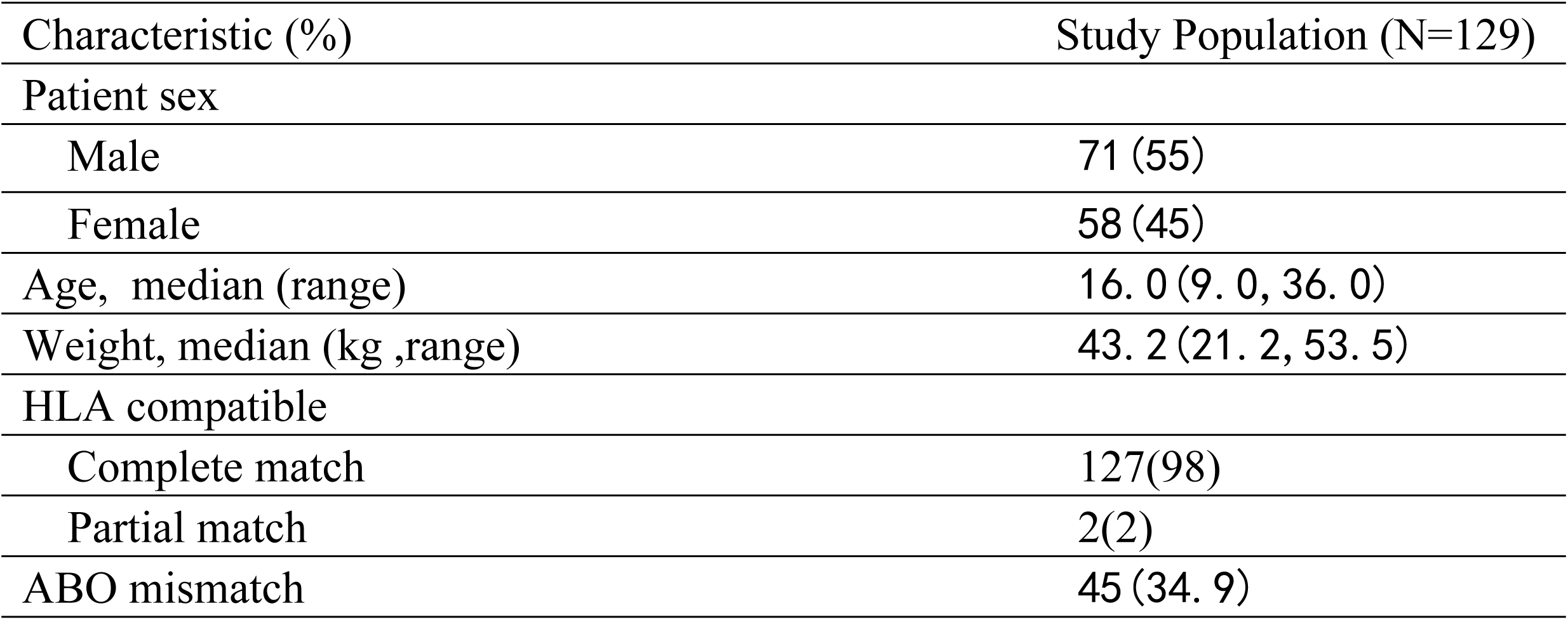
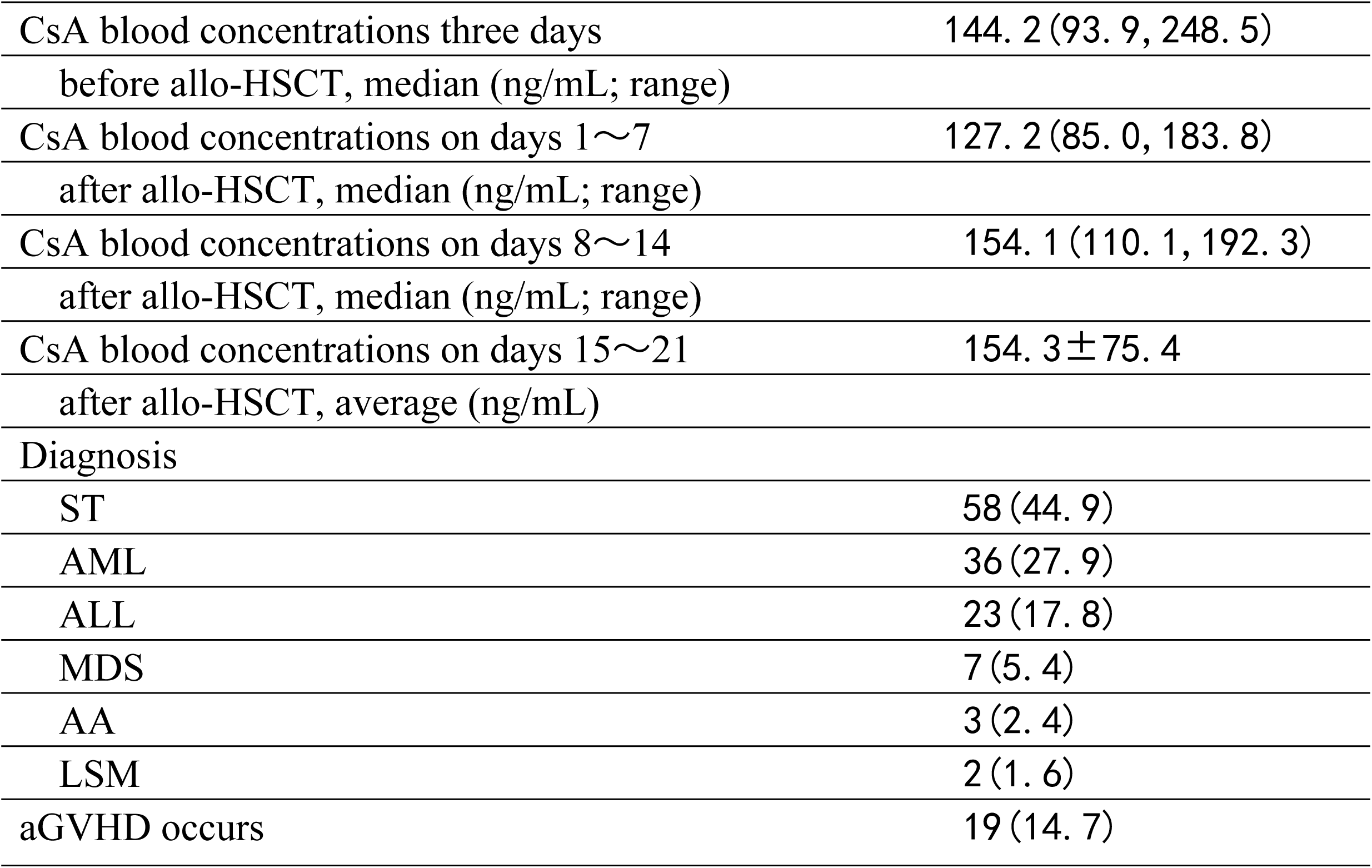
Study Population Characteristics [n, x̄±s; Md(P_25_,P_75_)]

| Table 1 | Study Population Characteristics | [n, $\bar{x}\pm s$ ; Md(P<sub>25</sub>,P<sub>75</sub>)] |
| --- | --- | --- |
| Characteristic (%) |  | Study Population (N=129) |
| Patient sex |  |  |
| Male |  | 71 (55) |
| Female |  | 58 (45) |
| Age, median (range) |  | 16. 0 (9. 0, 36. 0) |
| Weight, median (kg ,range) |  | 43. 2 (21. 2, 53. 5) |
| HLA compatible |  |  |
| Complete match |  | 127(98) |
| Partial match |  | 2(2) |
| ABO mismatch |  | 45 (34. 9) |

|  |  |
| --- | --- |
| CsA blood concentrations three days before allo-HSCT, median (ng/mL; range) | 144. 2 (93. 9, 248. 5) |
| CsA blood concentrations on days 1~7 after allo-HSCT, median (ng/mL; range) | 127. 2 (85. 0, 183. 8) |
| CsA blood concentrations on days 8~14 after allo-HSCT, median (ng/mL; range) | 154. 1 (110. 1, 192. 3) |
| CsA blood concentrations on days 15~21 after allo-HSCT, average (ng/mL) | 154. 3 ± 75. 4 |
| Diagnosis |  |
| ST | 58 (44. 9) |
| AML | 36 (27. 9) |
| ALL | 23 (17. 8) |
| MDS | 7 (5. 4) |
| AA | 3 (2. 4) |
| LSM | 2 (1. 6) |
| aGVHD occurs | 19 (14. 7) |

### Pretreatment programme

The pretreatment programme varies depending on according to the type of primary disease and the types of transplant, e.g. Busulfan (BU) + Cyclophosphamide (CY) + anti-thymocyte globulin (rabbit)(ATG)+Fludarabine(FLU); BU+CY +ATG)/Melphalan(Mel), and so on.

### Prevention strategies for aGVHD

Different GVHD prevention strategies vary depending on the source of the hematopoietic for stem cells. The sibling-matched regimen was based on CsA + Mycophenolic acid (MMF)+ short-term Methotrexate (MTX); in addition to the baseline unrelated or haploidentical transplantation regimen, ATG is administered at a dose of 2.5 mg/(kg·d) from day -4 to day -2.The initial dose of CsA is 3 mg/(kg·d), and the maintenance dose is adjusted based on changes in the patient’s clinical status and blood drug concentration.

### CsA blood concentration measurement

After 2 days of continuous CsA administration (intravenous or oral), 2 mL of peripheral venous blood were collected at a fixed time in the morning of each day or 30 minutes before oral drug administration for testing of blood concentration. The CsA concentrations in whole blood were determined using a Siemens Viva-E fully automated drug concentration analyser (enzyme immunoassay). Gently mix the blood sample thoroughly, add 100 μL blood sample and 300 μL sample pretreatment reagent to the 1.5 mL EP tube, then immediately cap and vortex for 20 seconds, let it stand at room temperature for 2 minutes, centrifuge at 14 000 rpm for 5 minutes and collect the supernatant for on-line testing. Blood concentrations of CsA should be measured 2 to 3 times per week, with the results from the first measurement of each week used for monitoring.

### CsA dose-normalized concentration

In order to eliminate the effect of the administered dose on the CsA blood concentration, the CsA blood concentration (C) must be divided by the administered dose of the day (D) to acquire the dose-normalized blood concentration (C/D) [ng-mL^-1^-mg^-1^]; C/D serves as a parameter reflecting the relationship between drug dosage and concentration ^[6]^. To exclude the effect of the route of administration on CsA blood concentrations, we administered CsA at a dose ratio of 1:2.5 when we switched the intravenous infusion of CsA to oral administration ^[7]^.

### Diagnosis of aGVHD

Classical aGVHD is generally defined as occurring up to 100d (+100d) post-transplantation with mainly inflammatory reactions in 3 organs: the skin, the gastrointestinal tract and the liver. aGVHD is diagnosed using a graded diagnostic scale based on criteria developed at the 1994 Seattle meeting ^[8]^.

### Statistical analysis

SPSS 26.0 software was used for statistical analysis. Normally distributed continuous data were expressed as mean ± standard deviation (x̄±s), while non-normally distributed continuous data were represented by Md (P_25_, P_75_). For one-way ANOVA, normally distributed continuous data underwent independent sample T-tests, whereas non-normally distributed data were subject to Mann-Whitney U tests. Categorical data was presented in terms of frequencies and percentages. Group comparisons were conducted using the Chi-square test, and statistically significant findings were incorporated into the multifactor analysis. The multifactor analysis employed a Logistic regression model. The significance level was set at α=0.05.

## Results

### aGVHD incidence

Among 129 patients followed up to 100 days post-transplantation, 19 cases of aGVHD occurred, yielding an incidence of 14.7%. The time to onset ranged from day 12 to day 59, with a mean of 31.4 days. Among the 19 cases of aGVHD, Grade I was observed in 10 patients, Grade III in 1 patient, and Grade IV in 8 patients, with one death recorded. The organs affected included the skin (11cases) and the gastrointestinal tract (8 cases). The early survival rate was 100% (10/10) in patients with grade I~II aGVHD and 88.9% (8/9) in patients with grade III~IV aGVHD, and the difference between them was not significant (*P*>0.05).

### Univariate analysis of aGVHD incidence

After allo-HSCT, patients were divided into the aGVHD group or the non-aGVHD group. Various factors including recipient age, donor-recipient gender relationship, HLA matching status, donor-recipient blood type compatibility, use ATG in conditioning regimen, presence chronic diseases before transplantation, bacterial and fungal infections before transplantation, EB virus infection before transplantation, post-transplant oral cavity infection, post-transplantation febrile neutropenia, infused MNCs cells, infused CD34 cells, time to neutrophil engraftment, time to platelet engraftment, pretreatment programme, CsA concentration 3 days before transplantation, CsA dose-normalized concentration 3 days before transplantation, CsA concentration on days 1~7 after transplantation, CsA dose-normalized concentration on days 1~7 after transplantation, CsA concentration on days 8~14 after transplantation, CsA dose-normalized concentration on days 8 ~ 14 after transplantation, CsA concentration on days 15 ~ 21 after transplantation, CsA dose-normalized concentration on days 15 ~ 21 after transplantation were analyzed as independent variables in a univariate analysis. The results demonstrated statistically significant differences in CsA concentration on days 15 ~ 21 after transplantation, bacterial infection before transplantation, fungal infection before transplantation, and the pretreatment programme, as detailed in Table 2.

**Table 2.** Univariate analysis results of aGVHD after allo-HSCT [x̄±s; Md(P_25_,P_75_)]

|  | aGVHD | Non-aGVHD | *P* |
| --- | --- | --- | --- |
|  | N=19 | N=110 |  |
| Age (years) , no. |  |  | 0. 624 |
| ≥ 16 | 11 | 57 |  |
| <16 | 8 | 53 |  |
| Donor-patient sex match, no. |  |  | 0. 985 |
| Matched | 9 | 54 |  |
| Male-female | 5 | 29 |  |
| Female-male | 5 | 27 |  |
| HLA type, no. |  |  | 0. 554 |
| 6/6 | 19 | 108 |  |
| 5/6 | 0 | 2 |  |
| ABO matched, no. |  |  | 0. 474 |
| Matched | 11 | 73 |  |
| Mismatched | 8 | 37 |  |
| Use ATG in conditioning regimen, no. |  |  | 0. 498 |
| Yes | 16 | 85 |  |
| No | 3 | 25 |  |
| Presence chronic diseases before transplantation, no. |  |  | 1. 0 |
| Yes | 3 | 14 |  |
| No | 16 | 94 |  |
| Bacterial infection before transplantation, no. |  |  | 0. 034 |
| Yes | 12 | 41 |  |
| No | 7 | 69 |  |
| Fungal infection before transplantation, no. |  |  | 0. 033 |
| Yes | 5 | 8 |  |
| No | 14 | 102 |  |
| EB virus infection before transplantation, no. |  |  | 0. 170 |
| Yes | 6 | 17 |  |
| No | 13 | 93 |  |
| Post-transplant oral cavity infections, no. |  |  | 0. 804 |
| Yes | 8 | 43 |  |
| No | 11 | 67 |  |
| Presence of febrile neutropenia after transplantation, no. |  |  | 0. 912 |
| Yes | 12 | 68 |  |
| No | 7 | 42 |  |
| Infused MNCs, X10<sup>8</sup>/kg (range) | 8. 91 ± 3. 88 | 10. 44 (7. 77, 13. 13) | 0. 220 |
| Infused CD34, X10<sup>8</sup>/kg (range) | 8. 63 ± 3. 67 | 6. 95 (4. 87, 10. 25) | 0. 165 |
| Time to neutrophil engraftment (days) | 13.0 (11.0, 14.0) | 11.5 (10.0, 13.0) | 0.139 |
| Time to platelet engraftment (days) | 13.0 (12.0, 15.0) | 13.0 (11.3, 15.8) | 0.986 |
| Pretreatment programme |  |  | <0.001 |
| Bu+Cy+ATG+Flu | 9 | 58 |  |
| Bu+Cy+ATG/Mel | 1 | 24 |  |
| Other | 9 | 28 |  |
| CsA concentration 3 days before transplantation, median (ng/mL; range) | 121.8 (70.7, 303.2) | 146.2 (95.5, 245.7) | 0.581 |
| CsA dose-normalized concentration 3 days before transplantation, median (ng/mL; range) | 70.4 (37.6, 104.4) | 67.6 (41.2, 102.1) | 0.737 |
| CsA concentration on days 1~7 after transplantation, median (ng/mL; range) | 121.8 (84.0, 176.9) | 128.3 (85.5, 185.5) | 0.655 |
| CsA dose-normalized concentration on days 1~7 after transplantation, average/median (ng/mL; range) | 72.7±31.4 | 76.9 (54.3, 116.6) | 0.313 |
| CsA concentration on days 8~14 after transplantation, average/median (ng/mL; range) | 158.0±50.0 | 150.4 (108.2, 195.3) | 0.883 |
| CsA dose-normalized concentration on days 8~14 after transplantation, median (ng/mL; range) | 79.2 (48.8, 116.4) | 79.0 (59.5, 116.4) | 0.675 |
| CsA concentration on days 15~21 after transplantation, average (ng/mL; range) | 196.3±87.6 | 147.2±71.1 | 0.010 |
| CsA dose-normalized concentration on days 15~21 after transplantation, average/median (ng/mL; range) | 110.7±58.6 | 82.5 (56.4, 145.0) | 0.452 |

In addition, further multifactorial logistic regression analyses were arried out considering CsA concentration, CsA dose-normalized concentration on days 15~21 after transplantation, bacterial and fungal infections before transplantation, pretreatment programme. The results indicated that CsA concentration, CsA dose-normalized concentration on days 15 ~ 21 after transplantation, bacterial infection before transplantation, pretreatment programme were independent risk factors for the aGVHD after allo-HSCT, as shown in Table 3.

**Table 3.** Multivariate analysis results of aGVHD after allo-HSCT.

|  | OR | 95%CI | *P* |
| --- | --- | --- | --- |
| CsA concentration on days 15 ~ 21 after transplantation | 0. 987 | 0. 978~ 0. 997 | 0. 011 |
| Bacterial infection before transplantation | 0. 255 | 0. 079~ 0. 823 | 0. 022 |
| Fungal infection before transplantation | 0. 467 | 0. 112~ 1. 946 | 0. 295 |
| Pretreatment programme | 0. 488 | 0. 252~0. 944 | 0. 033 |
| CsA dose-normalized concentration on days 15~21 after transplantation | 1. 015 | 1. 001~1. 029 | 0. 035 |

ROC curve analysis was performed to compare the two groups. The results demonstrated that using a cut-off value of 136.45 ng/ml for CsA concentration, and the predictive model based on CsA concentrationon on days 15 to 21 after transplantation achieved a sensitivity of 85.0%, specificity of 45.2%, and an area under the ROC curve (AUC) was 0.65 (*P*=0.034). The AUC under the ROC curve for Bacterial infection before transplantation was 0.333 (*P*=0.018). The AUC under the ROC curve for pretreatment programme was 0.653(*P*=0.031). The predictive model based on CsA dose-normalized concentrationon on days 15 to 21 after transplantation achieved a sensitivity of 65.0%, specificity of 60.6%, and the AUC under the ROC curve was 0.65 (*P*=0.345). The results are shown in Figure 1.

**Figure 1.**
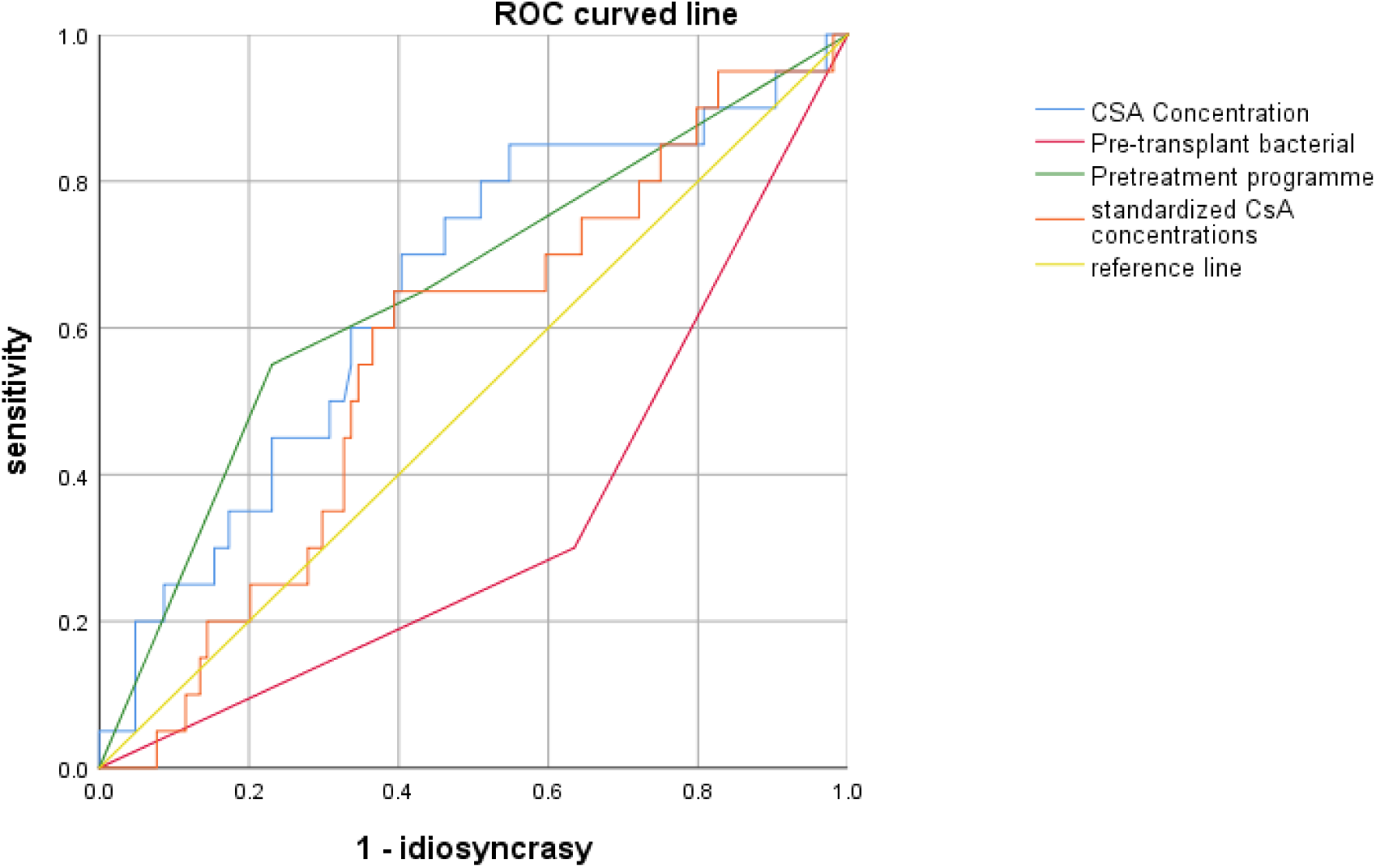
ROC curve analysis of the aGVHD and non-aGVHD groups after allo-HSCT

## Discussion

In this study, Research indicates that after allo-HSCT, aGVHD primarily affects the skin and gastrointestinal tract, with an incidence rate of 14.7%. Bacterial infection before transplantation, CsA concentrations and CsA dose-normalized concentrations on 15 ~ 21 after transplantation, as well as the pretreatment programme, have been identified as independent risk factors for aGVHD. The optimal critical concentration for CsA is determined to be 136.45 ng/ml.

The exact mechanisms underlying aGVHD are not yet fully understood. It is generally believed that the pathogenesis of aGVHD primarily involves the recognition and attack of host tissues by donor T cells, which occurs in three main phases: (1) Activation of the host’s antigen-presenting cells; (2) Activated host APCs, in conjunction with a variety of cytokines, activate donor T lymphocytes, leading to their activation, proliferation, and differentiation; (3) Activated donor T cells, along with cytokines, target organs, resulting in tissue damage and the clinical manifestations of aGVHD^[9]^. aGVHD has emerged as a major complication following allo-HSCT, posing significant risks to patients. The findings of this study indicate that 14.7% of patients experienced aGVHD, which is lower than the rates reported in the literature, which range from 30~50%^[2]^and 40~60%^[10]^. This may be attributed to the individualized administration of CsA, the combined use of novel immunosuppressants such as tacrolimus, mycophenolate mofetil, and sirolimus, as well as the implementation of rational conditioning regimens and a focus on maintaining a sterile environment. These measures have contributed to the gradual decline in the incidence of aGVHD.

CsA has a narrow therapeutic window, necessitating regular monitoring of drug concentrations. However, various factors can influence its plasma concentration levels, including red blood cell count, administration route, dietary habits, gastrointestinal function, drug interactions, as well as patient-specific variables such as sex and age ^[11]^.Therefore, fluctuations in CsA concentrations are common. Elevated CsA levels may lead to hepatotoxicity and nephrotoxicity, while subtherapeutic levels are associated with an increased risk of aGVHD^[12,13]^. Furthermore, low CsA concentrations particularly heighten the risk of developing grade II~IV aGVHD ^[13, 14]^. Consequently, an empirical dosing approach may not adequately meet patient needs. Early post-transplant monitoring of CsA plasma concentration remains the best strategy for preventing aGVHD ^[14]^. Although there have been reports ^[15]^ indicating the observation of peak concentrations to assess exposure to CsA, but the monitoring of trough concentrations remains the mainstream practice currently^[12,14]^. The relationship between CsA concentration levels and the occurrence of aGVHD remains a topic of debate, with differing opinions in the literature: The first perspective suggests that the trough concentration of CsA is related to both the incidence and severity of aGVHD ^[12、13、14]^. Maintaining a higher target concentration of CsA may be an effective strategy for preventing severe aGVHD^[14]^. Specifically, maintaining a trough concentration of CsA between 200 and 250 ng/mL can reduce the occurrence of aGVHD and its adverse effects^[16]^. Moreover, a target CsA concentration of 450 to 550 ng/mL has been associated with a lower incidence of aGVHD ^[17]^. The second perspective posits that the trough concentration of CsA is not related to the incidence of aGVHD ^[18 、 19]^. Early studies have shown that CsA can effectively reduce the severity of GVHD, it does not necessarily decrease its incidence ^[18]^. In fact, patients with severe grade III ~ IV aGVHD tend to have lower CsA concentration compared to those with grade I~II aGVHD, suggesting that lower CsA levels may predispose patients to more severe cases of aGVHD. However, by the third month post-transplant, no correlation has been found between aGVHD and CsA concentration ^[19]^. The conflicting conclusions regarding the relationship between CsA concentration and aGVHD may be attributed to several potential factors(: 1)This may be related to the various monitoring methods employed. While trough concentration monitoring is the standard practice, some studies have suggested that assessing CsA exposure at two or three time points post-transplant, particularly by monitoring the AUC ^[20]^.(2)The physiological characteristics, genetic backgrounds, and pathological conditions of different patients may lead to variations in drug metabolism, thereby influencing the relationship between CsA concentration and the occurrence of aGVHD.(3)The immune system response of patients varies due to individual differences, which may determine their reaction to CsA and susceptibility to GVHD. (4) The relationship between the trough concentration of CsA and aGVHD exhibits a “time window” characteristic ^[13, 14]^. P. Martin^[13]^ suggests that the relationship between the trough concentration of CsA and aGVHD features a “time-independent” period during the first two weeks post-transplant, as well as a “time-dependent” period leading up to the diagnosis of aGVHD. Although some reports indicate that low CsA concentrations in the first three weeks post-transplant are associated with a higher risk of aGVHD, however P. Martin emphasizes that the CsA concentration during the second week post-transplant is particularly critical. Malard F ^[14]^ found that the concentration of CsA during the first week post-transplant is significantly associated with the risk of severe grade II~IV aGVHD. In our study, we identified that the CsA concentration and dose-normalized concentration during the third week post-transplant serve as high-risk factors for the occurrence of aGVHD. Thus, the aforementioned discrepancies can be explained by variations in monitoring methods, immune differences among patients, and the differences in the “time window” associated with CsA concentration exposure. Research has revealed that the time window for CsA concentration falls within a “time-independent” period ^[13 、 14]^. In contrast, our study indicates that the time window for CsA concentration is within a “time-dependent” period. This suggests that there is a disparity in the relationship between CsA concentration and aGVHD. We hypothesize that this difference may be attributed to several factors: (1) The age of the population included in our study may be a contributing factor. The literature ^[13]^ focuses on children, while the study ^[14]^ pertains to elderly individuals. In contrast, our study population has a median age of 16 years. Age is an important factor that influences CsA concentration levels. (2) The sources of stem cells include matched siblings (MSD), mismatched relatives (MMRD), and unrelated donors (UD). Variations in the population included in different studies could contribute to differences in findings. Our study specifically focused on the parameter of CsA, thus primarily including populations from the MSD. The MMRD and UD predominantly received tacrolimus as part of their prophylactic regimen and were not included in this study. Additionally, the differences among various ethnic groups across different countries remain an unknown variable. (3) There are significant differences in the intensity of aGVHD prophylaxis among the study populations. (4) The pharmacokinetics of CsA are highly variable, and drug interactions within the study population can affect CsA concentration. For instance, the use of antifungal agents such as voriconazole may impact CsA levels. Therefore, there may be variations in the “time window” for CsA exposure. The key insight from this study is that, from the perspective of reducing healthcare costs for patients, for those undergoing HLA-matched transplants, the monitoring frequency of CsA can be adjusted to 1~2 times per week from the pre-transplant period through the first two weeks post-transplant, provided that the initial CsA concentration is within the target range. However, during the third week post-transplant, it may be beneficial to increase the monitoring frequency to every other day. This approach would allow for prompt adjustments in dosage in case of concentration fluctuations or abnormalities, facilitating timely attainment of target concentrations and potentially preventing the aGVHD. This strategy could be a feasible and effective measure. Although we have observed differences in the “time window” for CsA exposure, it is widely acknowledged that inadequate early exposure to CsA is a significant factor in the development of aGVHD. Therefore, closely monitoring CsA levels in the early post-transplant period, particularly during the first three weeks, may be an important strategy to mitigate this issue.

The consensus among Chinese experts (2020 edition) defines the effective trough concentration of CsA for sibling-matched transplantation as 150~250 ng/mL ^[21]^. China domestic studies have reported that the effective trough concentration of CsA is maintained at 200~250 ng/mL ^[16]^, or 200~400 ng/mL ^[22]^. Our study suggests that the occurrence of aGVHD is associated with CsA concentrations on days 15~21 after transplantation, with a cutoff value of 136.45 ng/mL during this period. This finding differs from previous studies, and we hypothesize that the reasons for this discrepancy may include: (1) The impact of detection methods on concentration levels ^[16]^. The domestic expert consensus ^[21]^ does not specify the detection methods employed. Currently, methods for measuring CsA concentration include enzyme amplification immunoassay, high-performance liquid chromatography, and liquid chromatography-mass spectrometry. Different detection methods may introduce variability and bias in the experimental data.(2)The impact of the pretreatment programme on reference values has been investigated. The study referenced ^[16]^ in utilized a modified BuCy regimen for conditioning and reached its conclusions accordingly. However, the domestic consensus ^[21]^ and the literature cited in ^[22]^ do not mention or include the pretreatment programme in their studies. In our research, we incorporated the pretreatment programme, and the results indicate that the pretreatment programme is an independent risk factor for the aGVHD.(3)There is a variation in the “time window” of CsA concentration exposure. The literature cited in ^[13]^ particularly focuses on the “time-independent” period, specifically the drug concentrations during the first two weeks post-transplant. However, our study reveals a strong correlation between the incidence of aGVHD and CsA concentrations during the third week post-transplant.(4)This may be related to the severity of the disease and age. One study suggests that children have lower CsA blood concentrations compared to adults, with target concentrations of 110 ng/ml for non-malignant conditions and 80 ng/ml for malignant diseases. In our study, the CsA concentrations were higher than those reported in the literature ^[13]^, primarily because the average age of our study population was 16 years, which is close to adults. Additionally, the target trough concentrations for CsA reported in the literature ^[16、21]^ are higher, mainly due to the lack of differentiation between adults and children.

Before allo-HSCT, bacterial infections are commonly observed. In a cohort of 129 patients studied, 53 patients (41.1%) experienced bacterial infections before transplantation, indicating a relatively high incidence rate. Patients typically undergo intensive chemotherapy or radiotherapy to eradicate malignant cells; however, these treatment modalities not only target cancer cells but also suppress normal hematopoietic cells. This suppression can lead to significant immunocompromise, particularly characterized by neutropenia. As a result, patients experience a decline in their immune defenses, particularly in their ability to ward off bacterial infections. Additionally, due to chemotherapy or other factors, patients may experience conditions such as oral mucositis or skin lesions, which create opportunities for bacterial invasion. Before allo-HSCT, fungal infections may also occur. In the cohort of 129 patients studied, 13 patients (10.0%) experienced fungal infections prior to transplantation, which is relatively lower compared to the incidence of bacterial infections. This discrepancy may be attributed to the fact that fungal infections typically require a greater degree of immunocompromise to develop. Fungal infections generally manifest in the context of prolonged immune suppression; thus, while patients may exhibit leukopenia during the early phase of immune suppression following chemotherapy, fungal infections are more commonly seen in later stages of sustained immune compromise, making them less prevalent prior to transplantation. Additionally, in some cases, hospitals may administer prophylactic antifungal agents prior to transplantation to further reduce the risk of fungal infections. Compared to bacteria, fungi generally have a slower replication rate, which can result in a delayed onset and manifestation of infection symptoms. This study found that the incidence of aGVHD following allo-HSCT was higher in patients with pre-transplant bacterial infections (63%) than in those with pre-transplant fungal infections (26%). The authors speculate that this may be due to the fact that bacterial infections trigger the release of a substantial amount of cytokines ^[23]^. These cytokines can influence the interactions between donor and recipient immune cells during the transplantation process, potentially leading to a stronger immune response. Bacterial infections can alter the host’s microenvironment, creating a state more conducive to inflammation and immune responses. This altered environment may facilitate the passive attack of donor cells during the transplantation process. In comparison, while fungal infections are also a significant type of infection, their impact on preoperative immune responses and microenvironment changes may differ from that of bacterial infections, potentially resulting in a lower incidence of aGVHD. The findings indicate that although pre-transplant fungal infections are not an independent risk factor for the development of aGVHD following transplantation, they can still contribute to other complications, such as systemic inflammatory response and immunosuppression. Therefore, effective infection management prior to transplantation is critically important.

This study found that the presence of bacterial infections before transplantation is an independent risk factor for the development of aGVHD post-transplant. Specifically, it was observed that bacterial infections can induce host cells to release pro-inflammatory cytokines such as interleukin-1 (IL-1), interleukin-6 (IL-6), and tumor necrosis factor-alpha (TNF-α). These inflammatory cytokines enhance the recognition of host MHC antigens by donor T lymphocytes and stimulate the production of interleukin-12 (IL-12) by host dendritic cells and macrophages. When inflammatory cytokines, such as IL-6, IL-8, and TNF-α, are highly expressed, the resulting “cytokine storm” is one of the significant contributors to the target organ damage associated with aGVHD ^[23]^. The gastrointestinal tract is not only a primary target organ for aGVHD but also serves as a significant amplifying system for this condition. Consequently, damage to the mucosal layer of the gastrointestinal tract can lead to an increased incidence of aGVHD. Studies have shown that when infectious factors are present, the body initiates an immune response against the pathogens. Concurrently, the extensive use of antibiotics to eliminate the gut microbiota may result in damage to the gastrointestinal mucosal epithelium, allowing bacterial degradation products, such as lipopolysaccharides, to enter the bloodstream. This chain reaction can lead to a “cytokine storm”. As a result, the probability of developing aGVHD in children with bacterial infections prior to transplantation is 2.63 times higher than in those without infections ^[24]^.Research has indicated that an imbalance in the gut microbiota is closely associated with the onset of post-operative aGVHD ^[2,3]^. The use of high-dose antimicrobial agents for bacterial infections prior to surgery can disrupt the gut microbiota and alter microbial metabolites, thereby compromising the balance of the intestinal microenvironment. This disruption significantly increases the risk of developing aGVHD ^[2]^. To mitigate the dysbiosis caused by factors such as infections, probiotics were administered from the time of transplantation until day 28 post-allo-HSCT, helping to improve the gut microecological environment and reduce the incidence of grade II~IV aGVHD ^[3]^. Therefore, patients with bacterial infections prior to surgery, timely intervention with probiotics and prebiotics following the use of antimicrobial agents may serve as an effective measure for the prevention and treatment of aGVHD.

This study demonstrated that the incidence of aGVHD following allo-HSCT was 14.7%, which is lower than the rates reported in previous literature ^[2, 5]^. This discrepancy may be related to the pretreatment programme, which is identified as an independent risk factor for the aGVHD. For patients with hematologic malignancies, such as leukemia or lymphoma, the pretreatment programme typically involves high-intensity chemotherapy or radiotherapy aimed at eliminating residual tumor cells in the body, thereby enhancing the success rate of transplantation.The pretreatment programme suppresses the patient’s immune system to reduce the risk of graft rejection. By putting the immune system into a “dormant” state, the process facilitates the survival and function of the graft. Additionally, conditioning eliminates unhealthy hematopoietic cells, thereby creating space in the bone marrow that allows for the growth and regeneration of the graft.The selection of pretreatment programme for allo-HSCT is influenced by various factors, including the patient’s age, type of disease, and performance status. Early myeloablative pretreatment programme included cyclophosphamide (Cy) combined with total body irradiation (TBI), busulfan (Bu) in combination with Cy, and modified BuCy (mBuCy) combined with anti-thymocyte globulin (ATG) ^[25]^. Subsequently, two major strategies for T-cell-depleted haplo-HSCT emerged: the Beijing protocol, which utilizes granulocyte colony-stimulating factor (G-CSF) and ATG to induce immune tolerance ^[21,26]^, and the Baltimore protocol, which employs post-transplantation cyclophosphamide (PTCy) to achieve similar outcomes ^[27,28]^. Both approaches have not only improved the success rate of haploidentical hematopoietic stem cell transplantation but also reduced the incidence of aGVHD. Early studies, such as those by Mehdizadeh et al. ^[29]^, indicated that myeloablative conditioning regimens were associated with a series of adverse reactions, including GVHD, which negatively impacted the overall life quality of patients. With the implementation of the post-transplantation cyclophosphamide (PTCy) protocol, a significant preventive effect against both severe aGVHD and chronic GVHD (cGVHD) has been observed, leading to its gradual incorporation into myeloablative pretreatment programme and HLA-matched transplants ^[30]^. However, reports have indicated that the use of PTCy alone in peripheral blood stem cell transplantation from HLA-matched sibling donors may increase the incidence and mortality of severe aGVHD ^[31]^. In contrast, employing a combined approach of PTCy and ATG has been shown to effectively control the incidence of aGVHD in patients with AML and MDS receiving HLA-matched peripheral blood hematopoietic stem cell transplants ^[32]^. For elderly patients aged 70 years and above with hematologic malignancies, the use of a melphalan-based reduced-intensity conditioning (RIC) regimen has shown favorable efficacy and acceptable toxicity. The cumulative incidence of grade II~IV aGVHD at 100 days post-transplantation was 37.7%, while the cumulative incidence of grade III~IV aGVHD was 18.9% ^[33]^. In the “Beijing protocol”, the use of rabbit anti-thymocyte globulin (rATG) is common in China, as it effectively depletes T cells in vivo, thereby reducing the incidence of aGVHD and significantly lowering the occurrence of cGVHD after HLA-matched transplantation ^[34]^. However, it is important to consider that the selection of rATG dosage should balance the risks of infection and aGVHD ^[35]^. Therefore, when formulating the optimal pretreatment programme, it is essential to consider individual differences, such as the recipient’s underlying condition, disease status, as well as the source of the donor and graft.

### Conclusions

In summary, for transplant patients using CsA for prophylaxis and treatment, in addition to individualizing the optimal pretreatment programme, pre-transplant bacterial infections may be managed with appropriate administration of prebiotics and probiotics to intervene in gut dysbiosis. Furthermore, post-transplantation, the monitoring frequency of CsA should be increased during the third week; if drug concentrations fluctuate or deviate from the normal range, the dose should be adjusted promptly to achieve the target concentration as early as possible, thereby preventing the occurrence of aGVHD.

## Data Availability

The datasets generated during and/or analysed during the current study are available from the corresponding author or Xi-Bin Wang on reasonable request.

## Abbreviations

allo-HSCT: allogeneic haematopoietic stem cell transplantation
aGVHD: Acute graft-versus-host disease
CsA: Cyclosporin
ST: Severe Thalassemia
AML: Acute Myeloid Leukemia
ALL: Acute Lymphoblastic Leukemia
MDS: Myelodysplastic Syndromes
AA: Aplastic anemia
LSM: Lymphatic System Malignancy
MTX: Methotrexate
MPA: Mycophenolic Acid
ATG: Antithymocyte Globulin
BU: Busulfan
CY: Cyclophosphamide
FLU: Fludarabine
Mel: Mefalan
HLA: Human leukocyte antigen
cGVHD: Chronic graft-versus-host disease

## Acknowledgments

We would like to thank all the patients participating in this study and all laboratory workers, nurses, and doctors who have assisted in the treatment in the stem cell transplantation.

## Authors’ contributions

T-TL designed the research; X-BW wrote the manuscript; and all authors provided the patients’ data and gave the final approval for the manuscript.

## Ethics approval and consent to participate

Informed consent was obtained from the patients and the children’s families, the study was conducted in accordance with the Declaration of Helsinki, and approved by the Ethics Committee of the First Affiliated Hospital of Guangxi Medical University (Ethics approval number: 2024-E748-01).

## TREND Statement Checklist

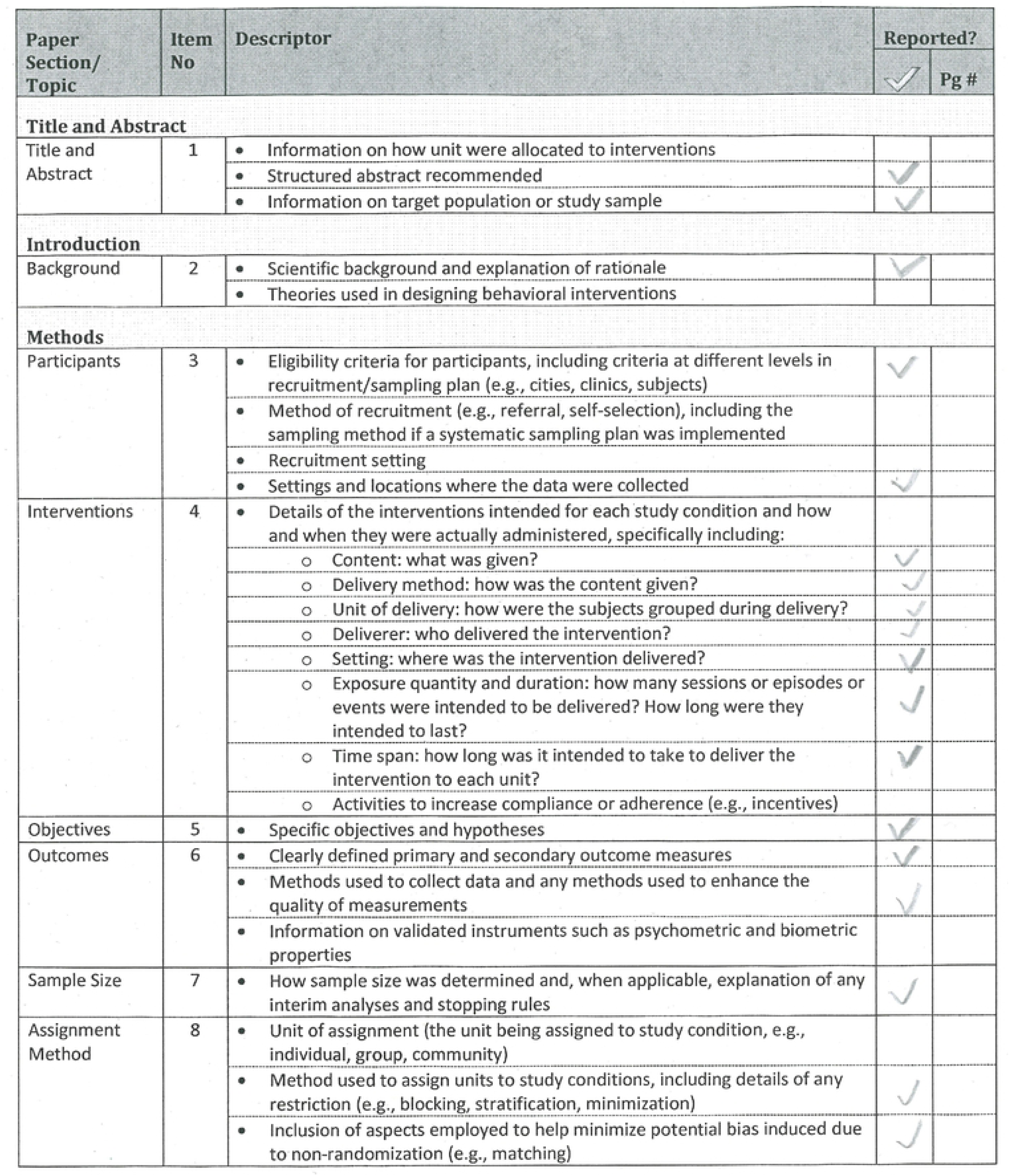
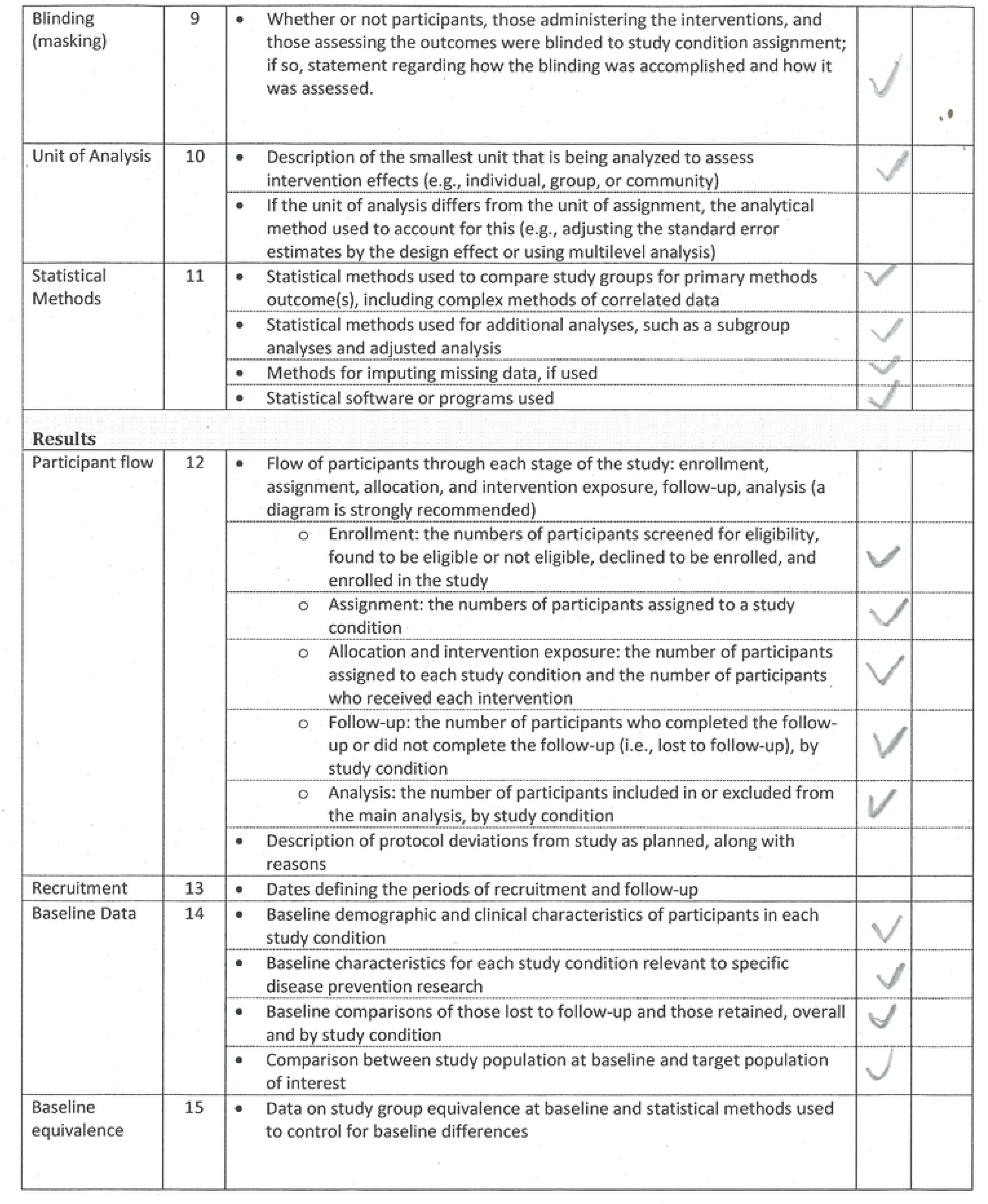
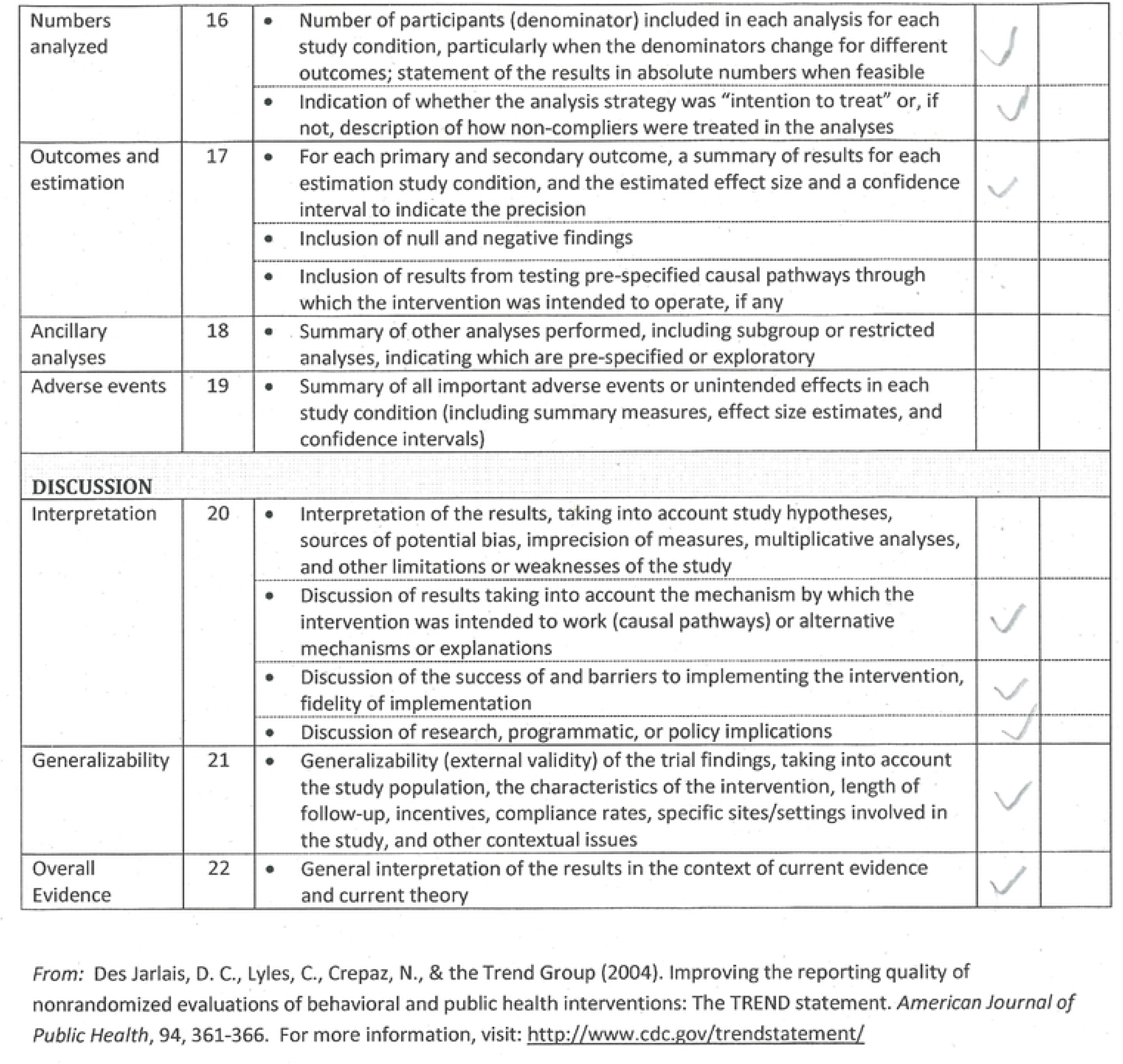

